# The Brain-Lung Immunotherapy Prognostic (BLIP) Score: A Novel Robust Tool for Prognostication in Non-Small Cell Lung Cancer Patients with Brain Metastases

**DOI:** 10.1101/2024.09.23.24314193

**Authors:** Marcus Skribek, Maria-Effrosyni Livanou, Ioannis Vathiotis, Viktor Strandman, Axel Thorell, Andreas Koulouris, Konstantinos Syrigos, Simon Ekman, Georgios Tsakonas

## Abstract

**Background:** Lung cancer remains the leading cause of cancer-related mortality, with brain metastases (BMs) significantly worsening prognosis and quality of life. The advent of immune checkpoint inhibitors (ICIs) has revolutionized the treatment landscape for non-small cell lung cancer (NSCLC). However, precise prognostic tools are essential to optimize clinical decision-making in this context.

**Methods:** The Brain-Lung Immunotherapy Prognostic (BLIP) score was developed based on a retrospective cohort of NSCLC patients treated with ICIs at Karolinska University Hospital, Sweden. Prognostic factors were identified using both univariate and multivariate Cox regression analyses. Internal validation was conducted using bootstrap resampling, penalized Cox regression, k-fold cross-validation, and receiver operating characteristics (ROC) analysis. External validation was performed using an independent cohort from Sotiria Thoracic Diseases Hospital of Athens, Greece.

**Results:** From a total cohort of 1844 patients screened across both study sites, 152 patients from Karolinska University Hospital and 116 from Sotiria Thoracic Diseases Hospital of Athens, Greece, were included in the final analysis. Key prognostic factors influencing outcomes included histology, actionable mutations, age at BM diagnosis, and the number of BMs. The BLIP score effectively stratified patients into two prognostic groups: “Good” and “Poor”, with a median overall survival (OS) of 15 and 7 months, respectively (hazard ratio [HR]: 0.4; *p* < 0.0001). External validation confirmed these findings, showing a significantly lower risk of death for the “Good” group compared to the “Poor” group (HR: 0.49; *p* = 0.0063). The model’s robust prognostic performance was confirmed with an area under the ROC curve of 0.87, highlighting its accuracy in predicting survival outcomes.

**Conclusion:** The BLIP score provides a reliable, validated prognostic tool for NSCLC patients with BMs undergoing ICI therapy. By integrating both molecular and clinical variables, it offers significant improvements over existing models. Prospective validation could further support its use in personalized treatment strategies, improving clinical outcomes and patient management.

**Highlights:**

- The BLIP score is a new prognostic tool for NSCLC.
- It focuses on patients with brain metastases undergoing immunotherapy.
- The score integrates clinical and molecular factors.
- Internal validation showed strong prognostic power and reliability.
- External validation confirmed effectiveness across diverse patient populations.
- Key factors include histology, actionable mutations, age, and brain metastases count.
- The score stratifies patients into “Good” and “Poor” groups.
- The BLIP score aids in personalized treatment decision-making.

## 1. Introduction

Lung cancer is the leading cause of cancer-related mortality, with non-small cell lung cancer (NSCLC) comprising most cases [1]. Brain metastases (BMs) are a frequent and severe complication in NSCLC patients, significantly affecting both prognosis and quality of life. Immune checkpoint inhibitors (ICIs) have revolutionized treatment for advanced lung cancer, improving survival outcomes [1]. However, BMs complicate treatment and prognostic evaluations, necessitating a personalized approach.

Approximately 25% of patients with metastatic malignancies develop BMs [2], with 20– 45% of lung cancer patients at risk [3,4]. BMs in NSCLC, particularly in adenocarcinoma and tumors harboring oncogenic driver alterations, are linked to poor prognosis [5]. ICIs, either combined with chemotherapy or as monotherapy, show promise for intracranial responses in NSCLC patients with BMs [6–10]. Nevertheless, management remains challenging due to patient heterogeneity, tumor variation, limited drug penetration into the brain, and resistance to prior therapies [11]. In this context, prognostic scores are crucial for individualized treatment approaches. By integrating clinical parameters, including patient-reported outcomes, biological markers, and disease characteristics, these tools facilitate treatment decisions, clinical trial stratification, and resource allocation.

The Recursive Partitioning Analysis (RPA) is a traditional clinical prognostic tool that categorizes patients based on age, Karnofsky performance status (KPS), primary tumor control, and presence of extracranial metastases [12]. However, it was developed using a relatively small, heterogenous population of patients who received whole-brain radiotherapy. The Graded Prognostic Assessment (GPA) incorporates age, KPS, extracranial metastases, and the number of metastases, which are validated prognostic factors [13,14]. Over time, the GPA has evolved to include disease-specific factors, enhancing its accuracy, but it does not incorporate molecular biomarkers [13]. Furthermore, neither the RPA nor the GPA takes into account the effects of ICIs. As personalized approaches advance, updates such as disease-specific GPA (DS-GPA) [15], Lung-molGPA [16], and NSCLC GPA [11] have incorporated oncogenic profiles and programmed death-ligand 1 (PD-L1) expression. However, PD-L1 exhibits significant intra- and intertumoral heterogeneity, resulting in inconsistent assessments, making it more predictive than prognostic [17–19]. The ALK-Brain Prognostic Index (ALK-BPI), developed for anaplastic lymphoma kinase (*ALK*)-positive NSCLC patients with BM treated with tyrosine kinase inhibitors, provides targeted insights but is limited to a specific patient population [20]. While several other clinical prognostic scores have been developed and validated [21–26], their utility for NSCLC patients with BMs requires further investigation.

Existing indices fail to capture the complexities introduced by ICIs in patients with BMs. This underscores the pressing need for a specialized prognostic tool tailored to this patient subset. The present study addresses this gap by developing and validating the Brain-Lung Immunotherapy Prognostic (BLIP) score, specifically designed for NSCLC patients with BMs.

## 2. Methods

### 2.1. Primary Patient Population and Study Design

This retrospective cohort study included patients diagnosed with NSCLC scheduled to receive ICIs at Karolinska University Hospital (KUH), Stockholm, Sweden, between July 2015 and August 2022. Patients were identified using the Cytodos software (CSAM Health, Norway), which manages the dosing, requisitioning, production, administration, and documentation of anticancer treatments.

Inclusion criteria were based on treatment received and diagnosis under ICD-10 code C34. Patients were excluded if they did not have primary NSCLC, had no BMs, did not receive ICIs during BM treatment, or were enrolled in prospective clinical trials.

Retrospective data was gathered from electronic medical records from KUH. The collected variables included sex, age, smoking status, histology, PD-L1 expression, genetic mutations, Eastern Cooperative Oncology Group Performance Status (ECOG PS) at BM diagnosis, number of BMs at BM diagnosis, BM response to prior therapy, location of metastases at lung cancer diagnosis, presence of extracranial metastases at BM diagnosis, BM classification (primary or secondary), symptomatic status of BM at BM diagnosis, details of treatments received, and outcome measures.

The study was approved by the regional ethical review board (approval number: 2020-02636) and adhered to the principles outlined in the Declaration of Helsinki. Due to the retrospective nature and pseudonymization of data, the ethical board waived the need for informed consent. Clinical stages were classified according to the 8^th^ edition of the TNM classification.

### 2.2. Statistical Analyses and Development of the BLIP Score

Categorical variables were analyzed using descriptive statistics, and statistical significance was set at *p* < 0.05 (two-sided test). Prognostic factors related to intracranial overall survival (OS) were identified using univariate and multivariate Cox proportional hazards regression to estimate hazard ratios (HR) and 95% confidence intervals (95% CI).

All clinical variables were initially assessed using univariate Cox regression. Variables showing statistical or clinical significance were included in the multivariate analysis. The backward stepwise elimination method was used to identify significant prognostic variables, which formed the basis for the BLIP score. Point allocations were based on regression coefficients and adjusted hazard ratios (aHRs).

The Kaplan-Meier method was used to assess the impact of the BLIP score on intracranial OS with binary outcomes, with the log-rank test comparing survival curves. Various cut-offs were tested for statistical significance, selecting the one with the highest prognostic value for the BLIP score. Intracranial OS was defined as the time from BM diagnosis to death, with living patients censored at the last follow-up. Importantly, patients had to receive ICIs while BMs were present to accurately capture the intracranial effect of ICIs, a unique criterion compared to similar studies. To ensure model robustness and assess multicollinearity among predictors, a variance inflation factor (VIF) analysis was conducted.

All statistical analyses were performed using RStudio (version 4.3.2) using the packages: readxl, survival, survminer, dplyr, writexl, car, caret, ggplot2, ROSE, glmnet, boot, pROC, tidyr and rmda (R Foundation for Statistical Computing, Vienna, Austria).

### 2.3. Internal Validation

The prognostic score was internally validated using multiple statistical methods. Data was split into training (60%) and testing (40%) sets to ensure robustness, providing a reasonable balance given the limited sample size while simulating real-world application. Kaplan-Meier survival curves were generated to visualize survival probabilities over time, with comparisons made using the log-rank test. Cox proportional hazards regression analyses were conducted to estimate HRs with 95% CI, assessing the model’s feasibility.

Model stability was tested using Efron-Gong bootstrap resampling, while penalized Cox regression (using LASSO) addressed overfitting and multicollinearity among predictors. The Partial Likelihood Deviance method was used for lambda (*λ*) selection in penalized regression, optimizing model parameters to minimize overfitting and maximize performance. Further validation involved 10-fold cross-validation, iteratively training and validating the model on different subsets to ensure performance was not dependent on a single train-test split. Together, these methods offer a comprehensive validation approach, with cross-validation focusing on the generalization aspect and bootstrap on the stability and variability of the model’s estimates. Calibration plots compared predicted probabilities with observed outcomes to assess accuracy. Performance metrics included the concordance index (c-index) for discriminative ability, Brier score to quantify prediction accuracy, and Receiver Operating Characteristics (ROC) curves with area under the curve (AUC) for sensitivity and specificity evaluation. Youden’s J statistic was used to determine the optimal threshold by maximizing the sum of sensitivity and specificity. Decision curve analysis evaluated the clinical utility of the prognostic model by assessing the net benefit across different threshold probabilities.

These steps provided a comprehensive internal validation of the prognostic score, ensuring its reliability and robustness in predicting clinical outcomes. Detailed explanations of the analyses are provided in the **Suppl. Data**.

### 2.4. External Validation

External validation of the BLIP score was performed using an independent cohort from Sotiria Thoracic Diseases Hospital of Athens, Greece. Data was collected retrospectively from medical records between December 2014 and July 2023. The inclusion and exclusion criteria were consistent with those used in the primary cohort at KUH. This approach ensured the robustness and effectiveness of the BLIP score across a different patient population and healthcare system. Patients were scored using the BLIP score, and survival outcomes were analyzed through Kaplan-Meier estimates, with log-rank tests used to compare survival curves. Cox regression analyses were employed to estimate HRs and 95% CI. All analyses were conducted locally. The study was approved by the regional ethical review board (approval number: 7450).

## 3. Results

### 3.1. Patient Characteristics

Out of 914 patients screened at KUH and 930 patients at Sotiria Thoracic Diseases Hospital of Athens, 152 and 116 patients, respectively, were included in the study (**Figure 1**). **Table 1** summarizes the patient characteristics. The primary cohort had a median follow-up of 17.5 months, with 58.6% female participants, whereas the validation cohort had a median follow up of 15.0 months and 77.6% male participants. The validation cohort had a higher proportion of current smokers (73.3% vs. 37.5%). Non-squamous carcinoma was predominant in both cohorts (91.4% in the primary cohort and 79.3% in the validation cohort). PD-L1 expression >50% was observed in 34.2% of the primary cohort and 21.6% of the validation cohort. Actionable mutations were present in 13.8% of the primary cohort and 6.0% of the validation cohort. The median age at BM diagnosis was 68 years in the primary cohort and 64 years in the validation cohort. ECOG PS 0–1 was seen in 83.5% (primary) and 67.2% (validation). Most patients had 1–3 BMs in both the primary (57.9%) and validation (81.9%) cohort. In the primary cohort, BM was diagnosed by CT in 40.1% and MRI in 59.9%, while in the validation cohort, CT was used in 28.4% and MRI in 70.7%. Pembrolizumab was the most used ICI in the primary cohort (72.4%), with nivolumab more common in the validation cohort (42.2%). Progressive disease was the primary reason for ICI discontinuation in both cohorts (75.0% primary, 78.4% validation). See **Suppl. Table 1** for more details.

**Figure 1:**
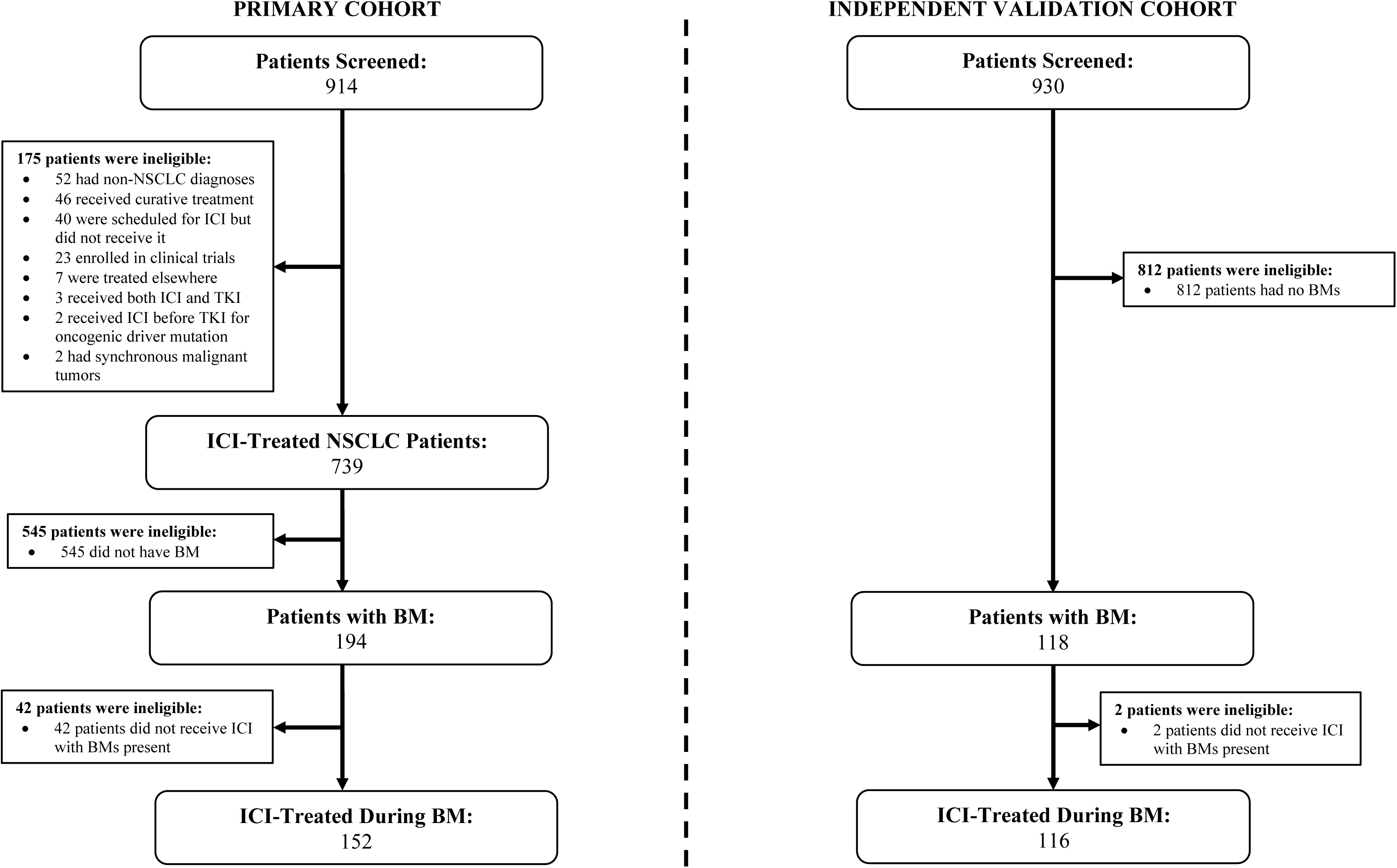
Flowchart of Study Design and Patient Population for NSCLC Patients Treated with ICIs Across Two Cohorts Abbreviations: NSCLC: non-small cell lung cancer; ICI: immune checkpoint inhibitor; TKI: tyrosine kinase inhibitor; BM: brain metastases.

**Table 1:**
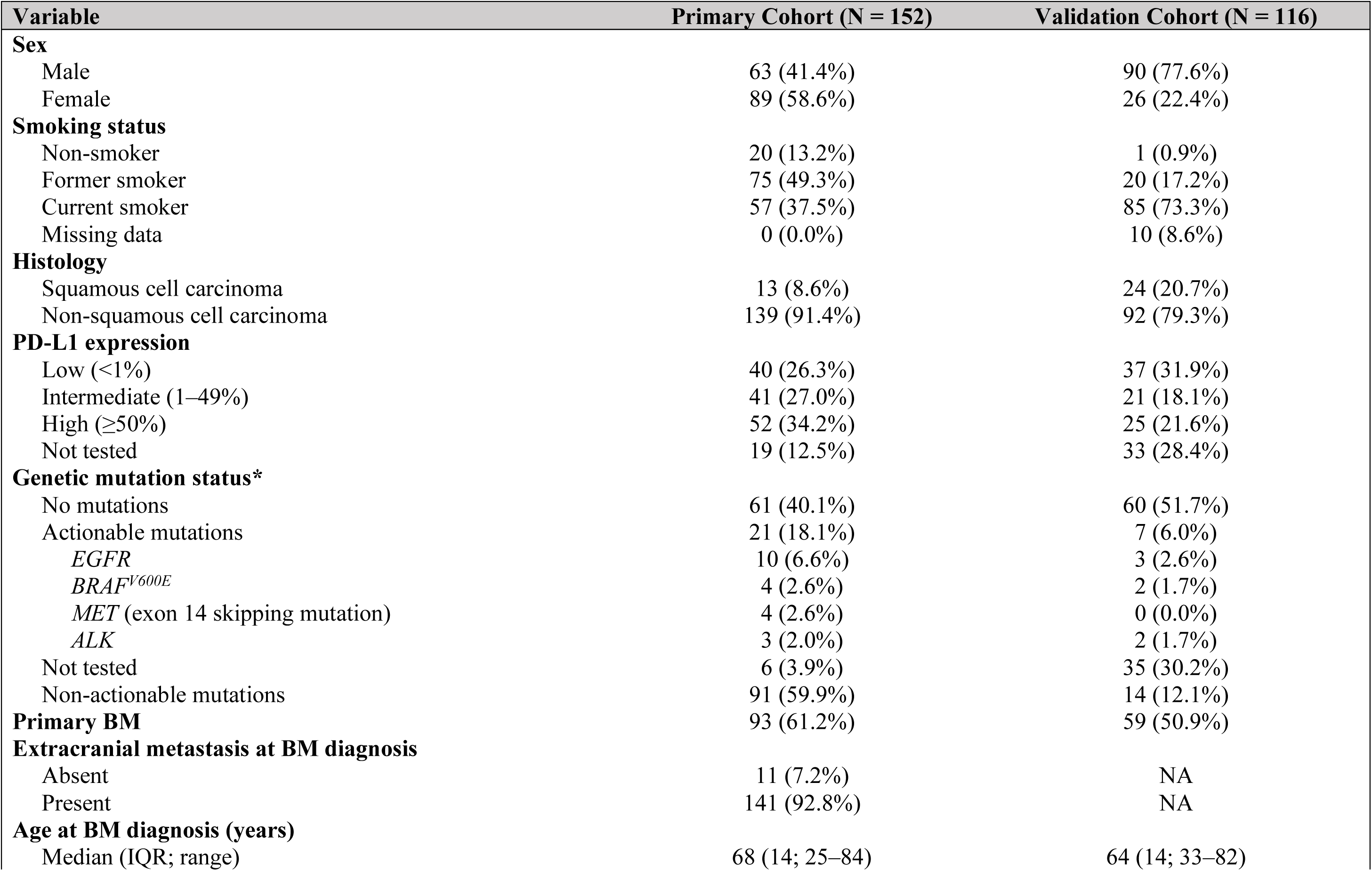
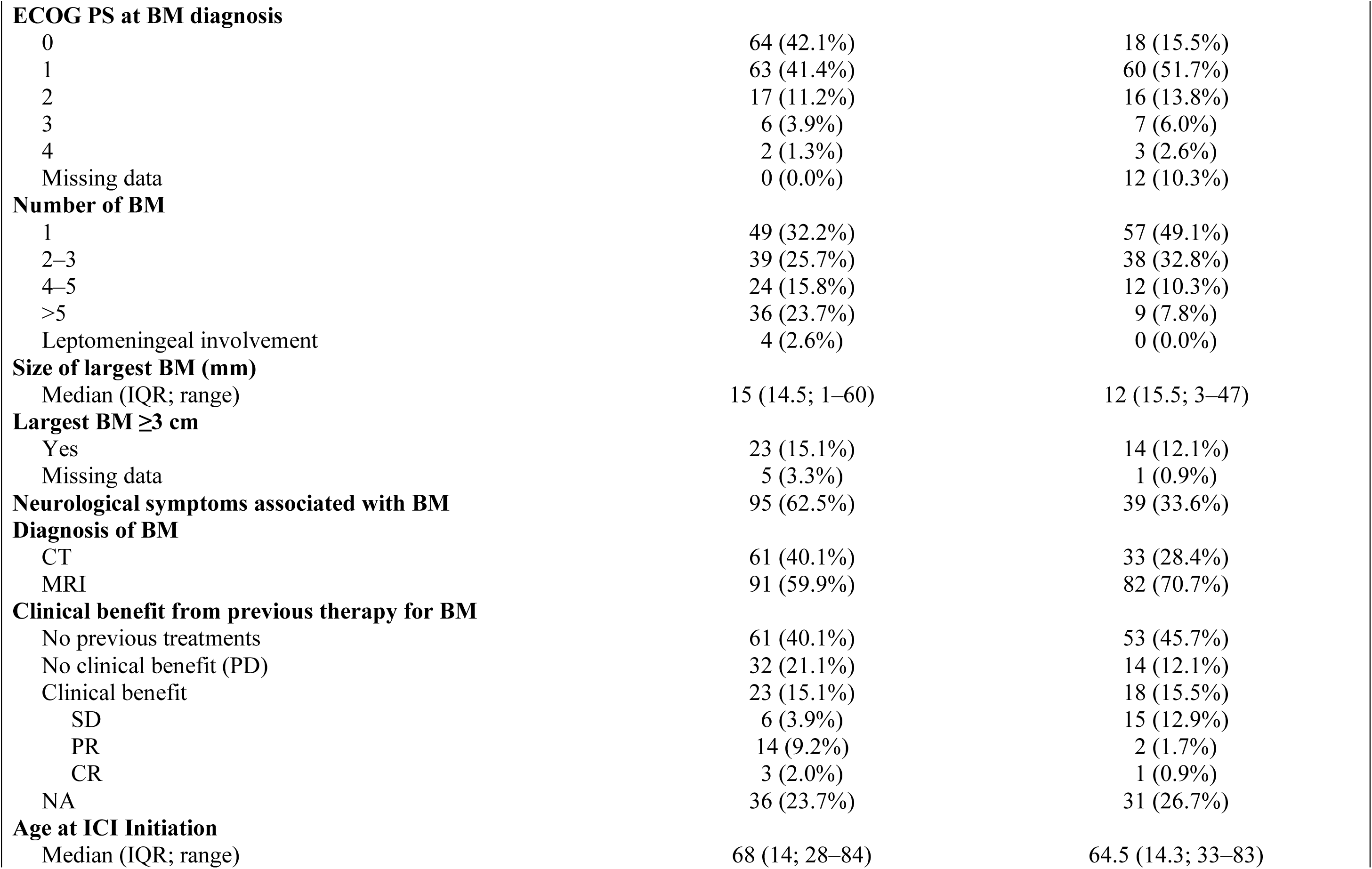
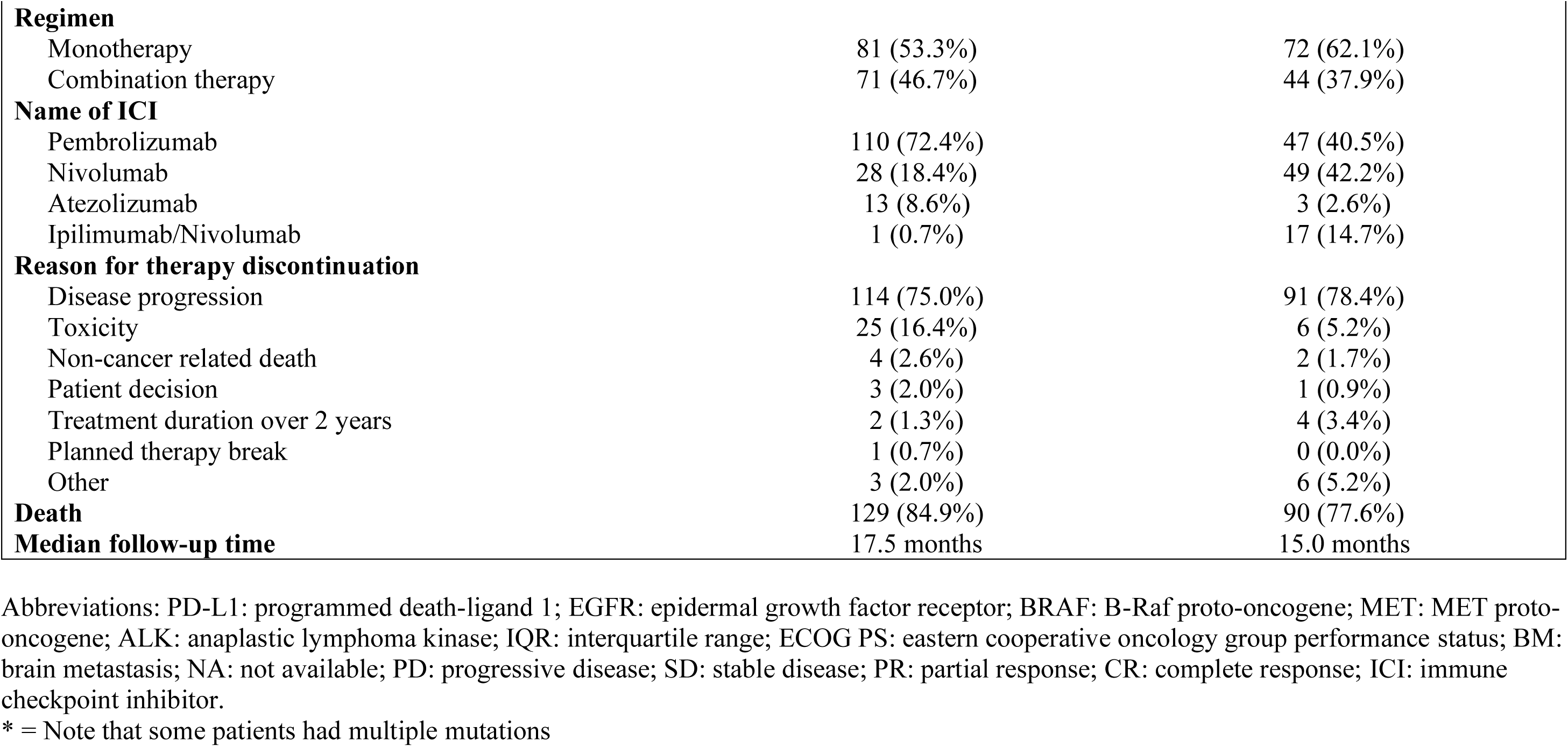
Patient Characteristics.

### 3.2. Clinical Factors Affecting the BLIP Score

Univariate analysis in the primary cohort identified factors associated with poor survival: squamous histology (HR = 3.17; 95% CI: 1.72–5.84; *p* < 0.001), thoracic metastasis (HR = 1.65; 95% CI: 1.15–2.38; *p* = 0.007), non-actionable mutations (HR = 0.56; 95% CI: 0.32–0.96; *p* = 0.034), age ≥65 (HR =1.77; 95% CI: 1.22–2.56; *p* = 0.003), higher ECOG PS (HR = 2.48; 95% CI: 1.20–5.13; *p* = 0.015), and more than three BMs (HR = 1.52; 95% CI: 1.07– 2.16; *p* = 0.021) (**Suppl. Table 2**). In the validation cohort, thoracic metastasis was not prognostic (HR = 0.85; 95% CI: 0.58–1.26; *p* = 0.42).

Multivariate analysis confirmed squamous histology (aHR = 3.77; 95% CI: 1.96–7.24; *p* < 0.001), non-actionable mutations (aHR = 0.52; 95% CI: 0.30–0.89; *p* = 0.017), older age (aHR = 1.57; 95% CI: 1.08–2.30; *p* = 0.018), and more than three BMs (aHR = 1.70; 95% CI: 1.17–2.46; *p* = 0.005) as significant predictors of intracranial OS. VIF values indicated no multicollinearity issues (**Table 2**).

**Table 2:**
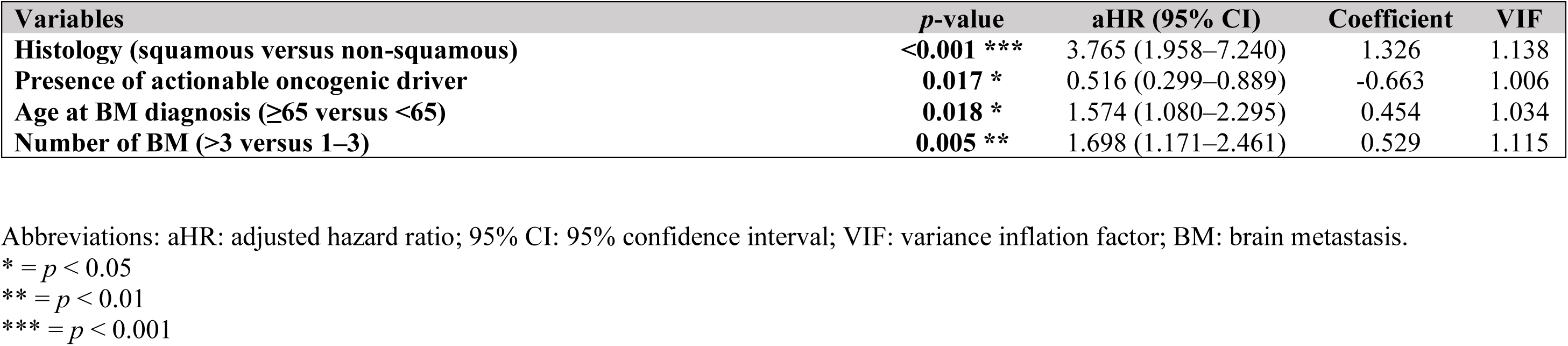
Multivariate Cox Proportional Hazards Regression Analysis of Variables Associated with Overall Survival Using Backward Stepwise Elimination.

| Variables | *p*-value | aHR (95% CI) | Coefficient | VIF |
| --- | --- | --- | --- | --- |
| Histology (squamous versus non-squamous) | <0.001 *** | 3.765 (1.958–7.240) | 1.326 | 1.138 |
| Presence of actionable oncogenic driver | 0.017 * | 0.516 (0.299–0.889) | -0.663 | 1.006 |
| Age at BM diagnosis (≥65 versus <65) | 0.018 * | 1.574 (1.080–2.295) | 0.454 | 1.034 |
| Number of BM (>3 versus 1–3) | 0.005 ** | 1.698 (1.171–2.461) | 0.529 | 1.115 |

Abbreviations: aHR: adjusted hazard ratio; 95% CI: 95% confidence interval; VIF: variance inflation factor; BM: brain metastasis.
\* = $p < 0.05$
\*\* = $p < 0.01$
\*\*\* = $p < 0.001$

### 3.3. Creation and Performance of the BLIP Score

Survival outcomes for prognostic factors are shown in **Table 3**. Median intracranial OS was 11.5 months (95% CI: 8–14) for the primary cohort and 12.0 months (95% CI: 10–15) for the validation cohort. Non-squamous histology had a median OS of 12.0 months (95% CI: 9–16) in the primary cohort versus 14.0 months (95% CI: 11–16) in the validation cohort. Squamous histology had a median OS of 5.0 months (95% CI: 2–not reached [NR]) in the primary cohort and 8.0 months (95% CI: 6–NR) in the validation cohort. Patients with actionable mutations had a median OS of 30.0 months (95% CI: 9–NR) in the primary cohort versus 16.0 months (95% CI: 9–NR) in the validation cohort. Points were allocated based on multivariate analysis, creating the BLIP score (**Table 4**). Kaplan-Meier estimates formed two prognostic groups: “Poor” (0–1 points) with a median OS of 7.0 months (95% CI: 4–10) and “Good” (2–4 points) with a median OS of 15 months (95% CI: 11–25). The HR was 0.40, indicating a 60% reduction in death risk for the “Good” group (**Figure 2A**). The difference between groups was highly significant (*p* < 0.0001).

**Figure 2:**
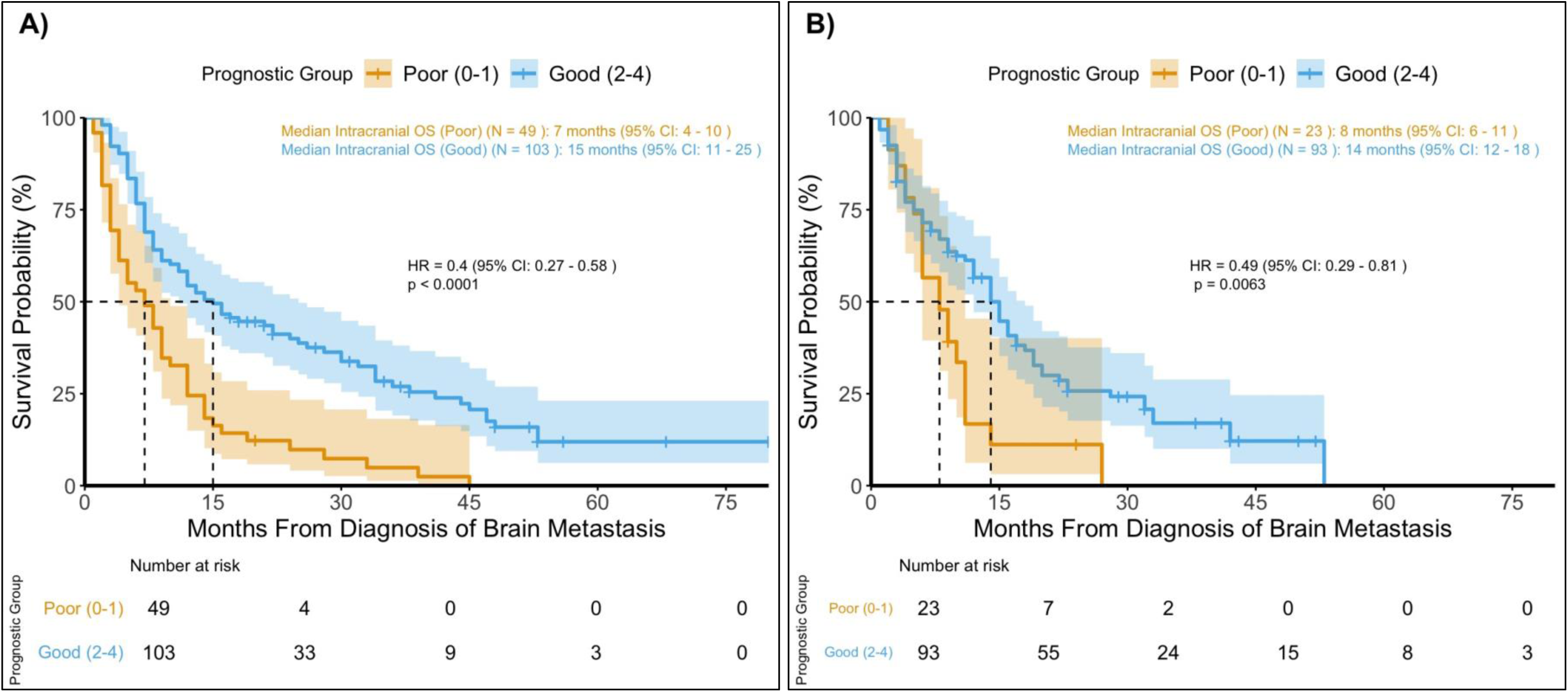
Kaplan-Meier Survival Curves Comparing Intracranial Overall Survival in A) Brain-Lung Immunotherapy Prognostic (BLIP) Groups and in the B) External Validation Cohort Abbreviations: OS: overall survival; 95% CI: 95% confidence interval; HR: hazard ratio.

**Table 3:** Survival Outcomes Associated with Prognostic Factors.

| Prognostic factor | Primary cohort (N = 152)<br>Median intracranial OS | Validation cohort (N = 116)<br>Median intracranial OS |
| --- | --- | --- |
| **Whole patient population** | 11.5 months (95% CI: 8–14) | 12.0 months (95% CI: 10–15) |
| **Histology** |  |  |
| Non-squamous cell carcinoma | 12.0 months (95% CI: 9–16) | 14.0 months (95% CI: 11–16) |
| Squamous cell carcinoma | 5.0 months (95% CI: 2–NR) | 8.0 months (95% CI: 6–NR) |
| **Presence of actionable oncogenic driver** |  |  |
| No or unknown | 10.0 months (95% CI: 8–14) | 12.0 months (95% CI: 9–15) |
| Yes | 30.0 months (95% CI: 9–NR) | 16.0 months (95% CI: 9–NR) |
| **Age at BM diagnosis** |  |  |
| <65 | 16.0 months (95% CI: 9–36) | 14.0 months (95% CI: 10–19) |
| ≥65 | 9.0 months (95% CI: 7–12) | 11.0 months (95% CI: 8–15) |
| **Number of BM** |  |  |
| 1–3 | 13.5 months (95% CI: 9–24) | 14.0 months (95% CI: 10–16) |
| >3 | 9.0 months (95% CI: 7–12) | 9.0 months (95% CI: 6–NR) |

Abbreviations: OS: overall survival; 95% CI: 95% confidence interval; NR: not reached; BM: brain metastasis.

**Table 4:**
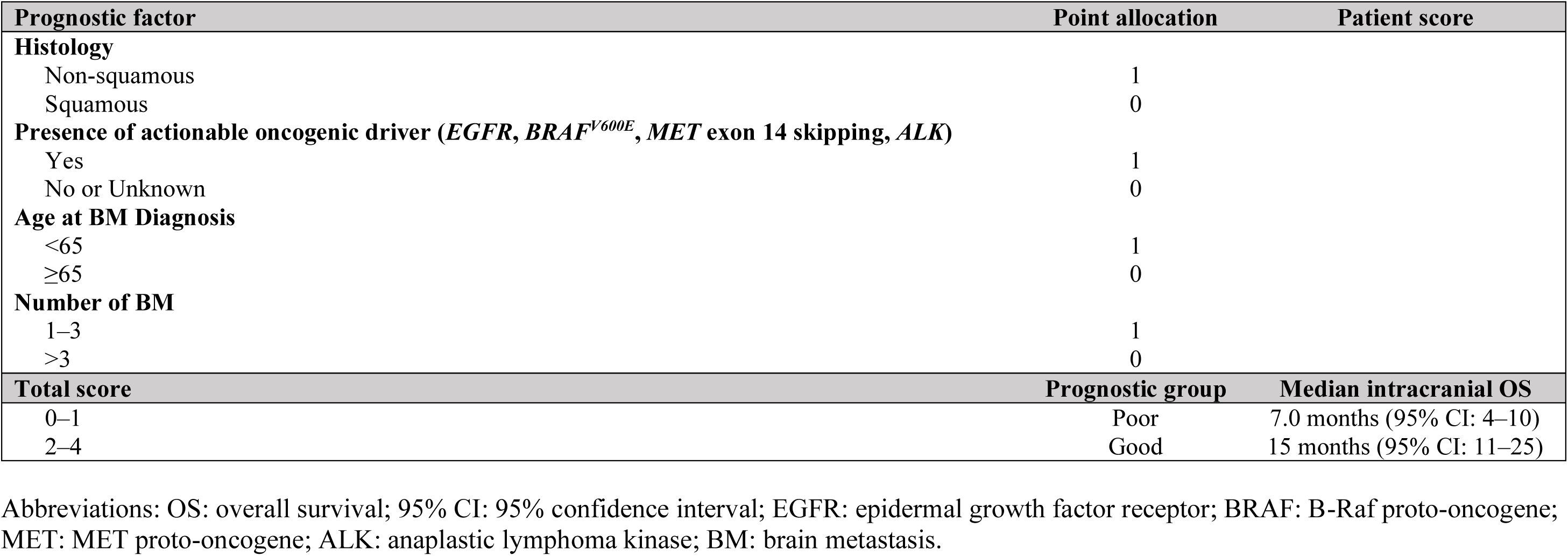
Calculation Worksheet for the Brain-Lung Immunotherapy Prognostic (BLIP) Score.

### 3.4. Internal Validation of the BLIP Score

The dataset was divided into training (60%, N = 92) and testing (40%, N = 60) cohorts (**Suppl. Figure 1**). Both cohorts showed significant survival differences between prognostic groups (training: *p* < 0.0001; testing: *p* = 0.003), demonstrating the BLIP score’s prognostic power.

Calibration plots (**Figure 3A**) showed strong agreement between predicted and observed survival probabilities, indicating excellent discrimination. The calibration curve closely followed the ideal line, indicating minimal bias. The Lasso regression model showed robust predictive performance and good calibration. Prognostic group stratification validated the model’s clinical utility. The ROSE package addressed class imbalance effectively. Partial likelihood deviance (**Suppl. Figure 2**) remained stable for *λ* values between −6 and −4.5. The optimal *λ* minimized deviance while balancing simplicity and accuracy. Predicted survival probabilities at 0, 1, 3, 6, 9, and 12 months for “Good” and “Poor” groups showed clear separation, supporting the model’s utility (**Figure 3B**).

**Figure 3:**
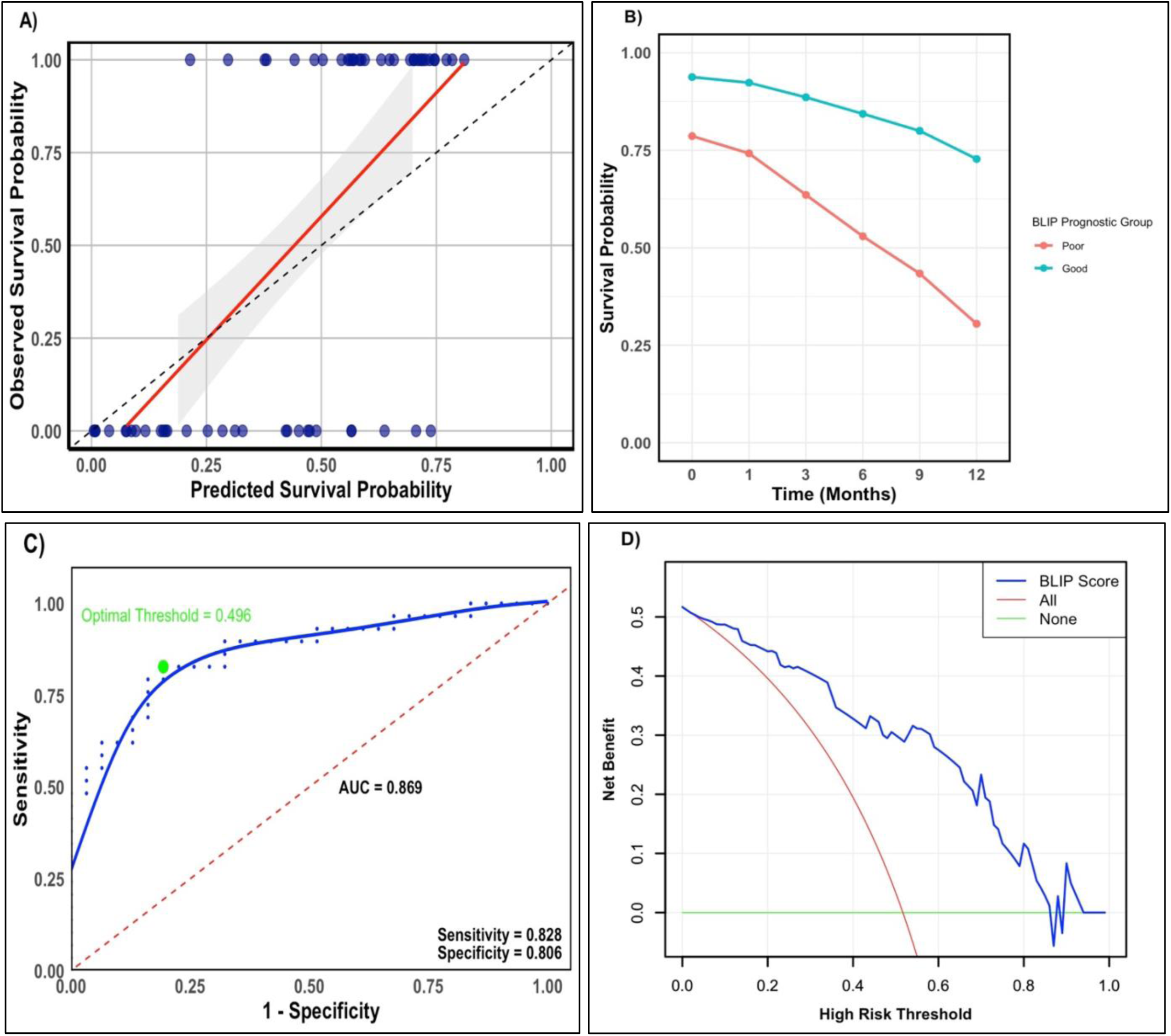
Internal Validation of the Brain-Lung Immunotherapy Prognostic (BLIP) Score: A) Calibration Plot for 12-Month Survival Probabilities, **B) Predicted Survival Probabilities, C) ROC Curve, and D) Decision Curve Analysis** Abbreviations: AUC: area under the curve; BLIP: Brain-Lung Immunotherapy Prognostic Score; ROC: receiver operating characteristics.

Performance metrics included a Brier score of 0.16, indicating accurate predictions, and a c-index of 0.630, showing moderate discrimination (**Suppl. Table 3**). The ROC curve with an AUC of 0.869 indicated excellent discrimination (**Figure 3C**). The optimal threshold (0.496) achieved 82.8% sensitivity and 80.6% specificity, enhancing clinical relevance.

Decision curve analysis (**Figure 3D**) showed the BLIP score’s higher net benefit across various thresholds, particularly lower ones. At a 0.2 threshold, the BLIP score’s net benefit was ∼0.45 compared to 0.20 for treat-all and 0 for treat-none strategies, confirming its utility in reducing overtreatment and improving decisions.

### 3.5. External Validation of the BLIP Score

In the validation cohort, Kaplan-Meier survival curves (**Figure 2B**) showed distinct outcomes. Patients in the “Poor” prognostic group (0–1 points) had a median intracranial OS of 8 months (95% CI: 6–11), while those in the “Good” prognostic group (2–4 points) had a median OS of 14 months (95% CI: 12–18). The hazard ratio for the “Good” group compared to the “Poor” group was 0.49 (95% CI: 0.29–0.81), significantly reducing the risk of death (*p* = 0.0063).

### 3.6. Sensitivity Analysis

A sensitivity analysis was conducted to identify the optimal model for evaluation, outlined in **Suppl. Table 3**. The analysis of regularization models revealed that Ridge, Lasso, and Elastic Net regressions each achieved an AUC of 0.8687, indicating their similar ability to rank predictions accurately. However, the Brier scores, which evaluate the accuracy of probabilistic predictions, differed slightly among the models. The Lasso regression model achieved the lowest Brier score of 0.1555, compared to Ridge (0.1574) and Elastic Net (0.1571). This suggests that Lasso regression provides slightly more accurate probability estimates. Despite identical AUC values, the superior calibration of the Lasso model, as evidenced by its lower Brier score, indicates it is the most suitable choice for this analysis.

## 4. Discussion

The BLIP score offers a novel and robust prognostic tool for NSCLC patients with BM undergoing ICI therapy. Unlike existing models, the BLIP score integrates molecular biomarkers with key clinical factors, capturing the unique dynamics introduced by ICIs. By incorporating variables such as histology, actionable oncogenic drivers, age at BM diagnosis, and the number of BMs, the BLIP score effectively stratifies patients into prognostic groups. Significant differences in intracranial OS were observed, with a median OS of 7 months for the “Poor” group and 15 months for the “Good” group. Internal validation confirmed strong discriminatory power, and calibration plots indicated minimal bias. Furthermore, decision curve analysis underscored the BLIP score’s clinical utility. External validation further solidified its robustness, showing a significant reduction in the risk of death in the “Good” group compared to the “Poor” group.

Existing prognostic tools, such as RPA [12], developed in patients treated with whole-brain radiotherapy, and GPA [13] guide survival predictions for BM but lack molecular data integration, which is now essential for personalized treatment approaches. These models, built on heterogenous populations, do not fully account for advancements like ICIs, which are pivotal in treating advanced NSCLC with BMs. While DS-GPA [15] incorporates clinical factors, it does not reflect the impact of ICIs, potentially leading to outdated prognostic estimates. More recent tools, such as Lung-molGPA [16] and NSCLC GPA [11], incorporate molecular markers like PD-L1, which significantly influence survival. However, reliance on this specific biomarker can lead to inconsistent assessments due to its variability [17–19]. In our study, PD-L1 was excluded from the multivariate analysis due to its lack of statistical significance, reflecting its uncertain prognostic value compared to its predictive role.

The survival differences between the BLIP score and other tools likely stem from differences in patient selection, treatment modalities, and prognostic factors. Unlike other scores, the BLIP score is tailored to a specific patient population, calculating OS from the date of BM diagnosis while receiving ICIs, making it highly relevant for current clinical practice. This specificity enables a more accurate understanding of BM in the context of modern treatment modalities, including ICIs.

Actionable and non-actionable alterations were separated to better reflect real-world practice. Although *KRAS* mutations were frequent, targeted therapies were unavailable during the study period. Although thoracic metastases at lung cancer diagnosis were significant in univariate analysis, they were excluded from the model due to unclear clinical relevance—a decision later validated by the external cohort. Additionally, the internal validation cohort’s wide 95% CI likely reflect its smaller size.

Oncogenic drivers are pivotal in determining prognosis and are included in the BLIP score. However, ICIs exhibit limited efficacy in patients with *EGFR*-mutant [27], *ALK*-positive [28] and *BRAF*-mutant [29] NSCLC. Notably, *BRAF*-mutant NSCLC, unlike melanoma, presents unique challenges, emphasizing the need to further investigate the sequencing of tyrosine kinase inhibitors and ICIs in this context [30]. Moreover, *KRAS* mutations complicate prognosis, as patients with these mutations may respond variably depending on the presence or absence of co-mutations (e.g., *KEAP1*, *STK11*, and TP53) [31–33]. Smoking further complicates this landscape by influencing genetic co-mutations and other genetic changes [34]. The survival benefit observed in our cohort may reflect real-world variations in the timing of tyrosine kinase inhibitor or ICI use, a practice that is yet to be standardized.

To our knowledge, the BLIP score is the first prognostic tool specifically developed for NSCLC patients with BMs undergoing ICI therapy. It integrates a comprehensive range of clinical and molecular variables, offering a tailored approach that enhances the accuracy of outcome predictions. The use of ECOG PS, rather than KPS, in the BLIP score reflects a variation in practice across different centers and clinical settings, with both scoring systems still in use depending on tradition and context. This adaptation aligns with contemporary clinical practices where ECOG PS has become more prevalent, particularly in ICI studies.

Both internal and external validations confirmed the BLIP score’s reliability, with external validation conducted in a diverse cohort with varying patient characteristics and healthcare systems. This extensive validation, to our knowledge in the largest study of its kind, ensures the score’s applicability across different clinical settings, highlighting its value in optimizing personalized treatment strategies for NSCLC patients with BM.

Despite its strengths, this study has several limitations. The retrospective design introduces potential biases and limits causal inference. The relatively small sample size may affect internal validity and increase the risk of Type II errors. Moreover, around 30% of patients in the validation cohort were not tested for oncogenic drivers, potentially influencing the findings. Finally, the limited inclusion of biomarkers may affect the precision of the prognostic model.

In conclusion, the BLIP score represents a significant advancement in the prognostication of NSCLC patients with BM undergoing ICI therapy, addressing critical gaps in existing models. Its rigorous validation, both internally and externally, confirms its reliability across diverse clinical settings. While the retrospective nature and relatively small sample size pose constraints, the BLIP score’s ability to accurately stratify patients underscores its potential to guide personalized treatment strategies. Future studies should aim to validate these findings in larger, multicentric (prospective) cohorts and explore the integration of additional biomarkers to further refine this prognostic tool.

## Data Availability

All data produced in the present study are available upon reasonable request to the authors.

## Acknowledgements

Gratitude is extended to Dr. Johan Zetterqvist, PhD at the department of Clinical Epidemiology at Karolinska Institutet for his invaluable assistance with the statistical methodology and analysis provided through the CLINICUM program. His expertise and guidance significantly contributed to the success of this research.

## Author Contributions

Conceptualization: M.S., S.E., G.T.; methodology and data acquisition: M.S., M.L., V.S., A.T., S.E., G.T.; formal analysis: M.S., G.T.; data interpretation: M.S., G.T.; writing—original draft preparation: M.S.; writing—review and editing: M.S., M.L., I.V., V.S., A.T., A.K., K.S., S.E., G.T.; visualization: M.S.; supervision: I.V., K.S., S.E., G.T.; resources and funding acquisition: M.S., S.E., G.T. All authors have read and agreed to the published version of the manuscript.

---

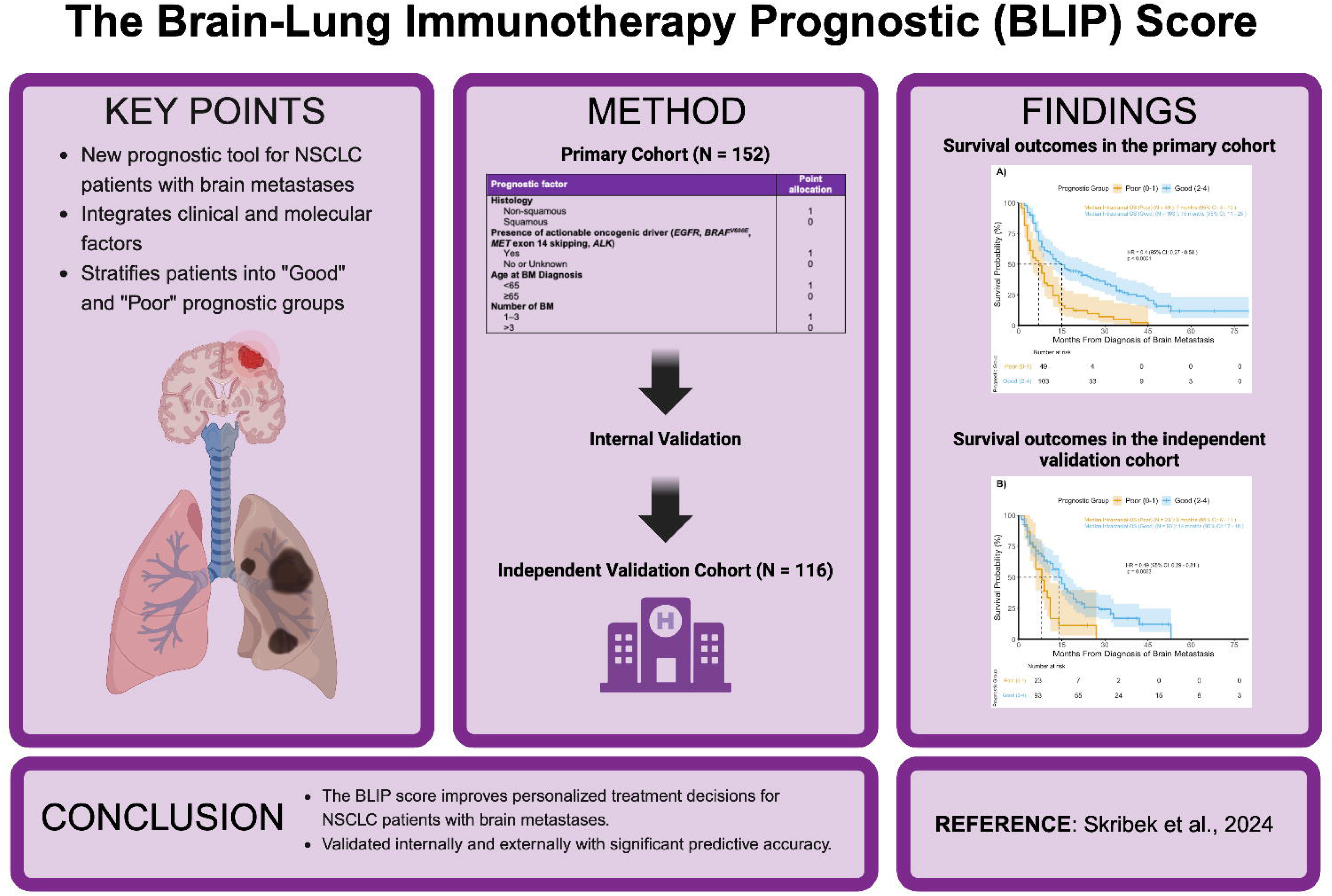

