## Supplementary Material for "The Brain-Lung Immunotherapy Prognostic (BLIP) Score: A Novel Robust Tool for Prognostication in Non-Small Cell Lung Cancer Patients with Brain Metastases"

| Variable | Primary Cohort (N = 152) | Validation Cohort (N = 116) |
|---|---|---|
| **Sex** | | |
| Male | 63 (41.4%) | 90 (77.6%) |
| Female | 89 (58.6%) | 26 (22.4%) |
| **Smoking status** | | |
| Non-smoker | 20 (13.2%) | 1 (0.9%) |
| Former smoker | 75 (49.3%) | 20 (17.2%) |
| Current smoker | 57 (37.5%) | 85 (73.3%) |
| Missing data | 0 (0.0%) | 10 (8.6%) |
| **Histology** | | |
| Adenocarcinoma | 126 (82.9%) | 79 (68.1%) |
| Squamous cell carcinoma | 13 (8.6%) | 24 (20.7%) |
| Other | 13 (8.6%) | 13 (11.2%) |
| NSCLC NOS/poorly differentiated | 10 (6.6%) | 4 (3.4%) |
| NSC NEC | 2 (1.3%) | 0 (0.0%) |
| Adenoid cystic | 1 (0.7%) | 0 (0.0%) |
| Large cell neuroendocrine carcinoma | 0 (0.0%) | 6 (5.2%) |
| Combined histology | 0 (0.0%) | 2 (1.7%) |
| Sarcomatoid | 0 (0.0%) | 1 (0.9%) |
| **Stage at lung cancer diagnosis** | | |
| 1 | 4 (2.6%) | 2 (1.7%) |
| 2 | 5 (3.3%) | 5 (4.3%) |
| 3 | 13 (8.6%) | 22 (19.0%) |
| 4 | 130 (85.5%) | 87 (75.0%) |
| **Location of metastases at lung cancer diagnosis** | | |
| Thoracic | 60 (39.5%) | 46 (39.7%) |
| Lymph node | 122 (80.2%) | 90 (77.6%) |
| Brain | 93 (61.2%) | 57 (49.1%) |
| Liver | 27 (17.8%) | 12 (10.3%) |
| Bone | 39 (25.7%) | 26 (22.4%) |
| Adrenal gland | 21 (13.8%) | 19 (16.4%) |
| None | 6 (3.9%) | 11 (9.5%) |

| | | |
|---|---|---|
| Other | 18 (11.8%) | 35 (30.2%) |
| Subcutaneous | 5 (3.3%) | 1 (0.9%) |
| Muscle | 2 (1.3%) | 3 (2.6%) |
| Renal | 5 (3.3%) | 2 (1.7%) |
| Colon | 3 (2.0%) | 1 (0.9%) |
| Pancreas | 2 (1.3%) | 0 (0.0%) |
| Ocular | 1 (0.7%) | 0 (0.0%) |
| Extrathoracic lymph node | 0 (0.0%) | 21 (18.1%) |
| Soft tissue | 0 (0.0%) | 5 (4.3%) |
| Spleen | 0 (0.0%) | 2 (1.7%) |
| **PD-L1 expression** | | |
| Low (<1%) | 40 (26.3%) | 37 (31.9%) |
| Intermediate (1–49%) | 41 (27.0%) | 21 (18.1%) |
| High (≥50%) | 52 (34.2%) | 25 (21.6%) |
| Not analyzed | 19 (12.5%) | 33 (28.4%) |
| **Genetic mutation status*** | | |
| No mutation | 61 (40.1%) | 60 (51.7%) |
| Actionable mutations | 21 (18.1%) | 7 (6.0%) |
| *EGFR* | 10 (6.6%) | 3 (2.6%) |
| *BRAF*$^{V600E}$ | 4 (2.6%) | 2 (1.7%) |
| *MET* (exon 14 skipping mutation) | 4 (2.6%) | 0 (0.0%) |
| *ALK* | 3 (2.0%) | 2 (1.7%) |
| Not tested | 6 (3.9%) | 35 (30.2%) |
| Non-actionable mutations | 91 (59.9%) | 14 (12.1%) |
| *KRAS* | 49 (32.2%) | 11 (9.5%) |
| *TP53* | 8 (5.3%) | 4 (3.4%) |
| *MET* (other than exon 14 skipping mutations) | 7 (4.6%) | 0 (0.0%) |
| *PIK3CA* | 3 (2.0%) | 1 (0.9%) |
| *PTEN* | 2 (1.3%) | 0 (0.0%) |
| *BRAF* (other than V600E) | 2 (1.3%) | 0 (0.0%) |
| *HER2* | 2 (1.3%) | 1 (0.9%) |
| *CTNNB1* | 1 (0.7%) | 1 (0.9%) |

| | | |
|---|---|---|
| *MYC*-amplification | 1 (0.7%) | 0 (0.0%) |
| *CEBP* | 1 (0.7%) | 0 (0.0%) |
| *ALK* exon 23 | 1 (0.7%) | 0 (0.0%) |
| *MDM4* | 1 (0.7%) | 0 (0.0%) |
| *C-KIT* | 1 (0.7%) | 0 (0.0%) |
| *MYB-NFIB* | 1 (0.7%) | 0 (0.0%) |
| *CDK4* | 1 (0.7%) | 0 (0.0%) |
| *CDK6* | 1 (0.7%) | 0 (0.0%) |
| *FOXA1* | 1 (0.7%) | 0 (0.0%) |
| *KIF5B* | 1 (0.7%) | 0 (0.0%) |
| *RET* | 1 (0.7%) | 1 (0.9%) |
| *MAP2K1* | 1 (0.7%) | 0 (0.0%) |
| *TERT* | 1 (0.7%) | 0 (0.0%) |
| *NRAS* | 1 (0.7%) | 0 (0.0%) |
| *SMARCA4* | 1 (0.7%) | 0 (0.0%) |
| *DICER1* | 1 (0.7%) | 0 (0.0%) |
| *NF1* | 1 (0.7%) | 0 (0.0%) |
| *MODY1* | 0 (0.0%) | 1 (0.9%) |
| *RB1* | 0 (0.0%) | 1 (0.9%) |
| *AKT1* | 0 (0.0%) | 1 (0.9%) |
| *NTRK2* | 0 (0.0%) | 1 (0.9%) |
| *RET* | 0 (0.0%) | 1 (0.9%) |
| **Primary BM** | 93 (61.2%) | 59 (50.9%) |
| **Extracranial metastasis at BM diagnosis** | | |
| Absent | 11 (7.2%) | NA |
| Present | 141 (92.8%) | NA |
| **Age at BM diagnosis (years)** | | |
| Median (IQR; range) | 68 (14; 25–84) | 64 (14; 33–82) |
| **ECOG PS at BM diagnosis** | | |
| 0 | 64 (42.1%) | 18 (15.5%) |
| 1 | 63 (41.4%) | 60 (51.7%) |
| 2 | 17 (11.2%) | 16 (13.8%) |

| | | |
|---|---|---|
| 3 | 6 (3.9%) | 7 (6.0%) |
| 4 | 2 (1.3%) | 3 (2.6%) |
| Missing data | 0 (0.0%) | 12 (10.3%) |
| **Number of BM** | | |
| 1 | 49 (32.2%) | 57 (49.1%) |
| 2–3 | 39 (25.7%) | 38 (32.8%) |
| 4–5 | 24 (15.8%) | 12 (10.3%) |
| >5 | 36 (23.7%) | 9 (7.8%) |
| Leptomeningeal involvement | 4 (2.6%) | 0 (0.0%) |
| **Size of largest BM (mm)** | | |
| Median (IQR; range) | 15 (14.5; 1–60) | 12 (15.5; 3–47) |
| **Largest BM ≥3 cm** | | |
| Yes | 23 (15.1%) | 14 (12.1%) |
| Missing data | 5 (3.3%) | 1 (0.9%) |
| **Neurological symptoms associated with BM** | 95 (62.5%) | 39 (33.6%) |
| **Diagnosis of BM** | | |
| CT | 61 (40.1%) | 33 (28.4%) |
| MRI | 91 (59.9%) | 82 (70.7%) |
| **Clinical benefit to previous line of therapy for BM** | | |
| No previous treatments | 61 (40.1%) | 53 (45.7%) |
| No clinical benefit (PD) | 32 (21.1%) | 14 (12.1%) |
| Clinical benefit | 23 (15.1%) | 18 (15.5%) |
| SD | 6 (3.9%) | 15 (12.9%) |
| PR | 14 (9.2%) | 2 (1.7%) |
| CR | 3 (2.0%) | 1 (0.9%) |
| NA | 36 (23.7%) | 31 (26.7%) |
| **Age at ICI Initiation** | | |
| Median (IQR; range) | 68 (14; 28–84) | 64.5 (14.3; 33–83) |
| **Name of ICI** | | |
| Pembrolizumab | 110 (72.4%) | 47 (40.5%) |
| Nivolumab | 28 (18.4%) | 49 (42.2%) |
| Atezolizumab | 13 (8.6%) | 3 (2.6%) |

| | | |
|---|---|---|
| Ipilimumab/Nivolumab | 1 (0.7%) | 17 (14.7%) |
| **Regimen** | | |
| Monotherapy | 81 (53.3%) | 72 (62.1%) |
| Pembrolizumab/Pemetrexed/Platinum | 60 (39.5%) | 19 (16.4%) |
| Pembrolizumab/(Nab)Paclitaxel/Platinum | 4 (2.6%) | 5 (4.3%) |
| Pembrolizumab/Pemetrexed | 4 (2.6%) | 0 (0.0%) |
| Ipilimumab/Nivolumab/Platinum/Paclitaxel | 1 (0.7%) | 4 (3.4%) |
| Ipilimumab/Nivolumab/Platinum/Pemetrexed | 0 (0.0%) | 13 (11.2%) |
| Atezolizumab/(Nab)Paclitaxel/Platinum/Bevacizumab | 1 (0.7%) | 1 (0.9%) |
| Atezolizumab/Platinum/Etoposide | 1 (0.7%) | 2 (1.7%) |
| **Line of ICI therapy in metastatic lung cancer** | | |
| Median (IQR; range) | 1 (1; 1–6) | 2 (1; 1–5) |
| **Number of cycles of ICI** | | |
| Median (IQR; range) | 6 (7; 1–35) | 4 (7; 1–50) |
| **Duration of ICI (days)** | | |
| Median (IQR; range) | 92.5 (167; 1–785) | 70.5 (187; 1–802) |
| **Reason for therapy discontinuation** | | |
| Disease progression | 114 (75.0%) | 91 (78.4%) |
| Toxicity | 25 (16.4%) | 6 (5.2%) |
| Non-cancer related death | 4 (2.6%) | 2 (1.7%) |
| Patient decision | 3 (2.0%) | 1 (0.9%) |
| Treatment duration over 2 years | 2 (1.3%) | 4 (3.4%) |
| Planned therapy break | 1 (0.7%) | 0 (0.0%) |
| Other | 3 (2.0%) | 6 (5.2%) |
| **Death** | 129 (84.9%) | 90 (77.6%) |
| **Median follow-up time** | 17.5 months | 15.0 months |

**Suppl. Table 1: Detailed Patient Characteristics**

Abbreviations: NSCLC NOS: non-small cell lung cancer not otherwise specified; NSC NEC: non-small cell neuroendocrine carcinoma; PD-L1: programmed death-ligand 1; EGFR: epidermal growth factor receptor; BRAF: B-Raf proto-oncogene; MET: MET proto-oncogene; ALK: anaplastic lymphoma kinase; KRAS: Kirsten rat sarcoma viral oncogene homolog; TP53: tumor protein p53; PIK3CA: phosphatidylinositol-4,5-bisphosphate 3-kinase catalytic subunit alpha; PTEN: phosphatase and tensin homolog; HER2: human epidermal growth factor receptor 2; CTNNB1: catenin beta 1; MYC: MYC proto-oncogene; CEBP: CCAAT/enhancer binding protein; MDM4 proto-oncogene; C-KIT: KIT proto-oncogene; MYB-NFIB: MYB proto-oncogene, NFIB gene fusion; CDK4: cyclin-dependent kinase 4; CDK6: cyclin-dependent kinase 6; FOXA1: forkhead box a1; KIF5B: kinesin family member 5b; RET: Ret proto-oncogene; MAP2K1: mitogen-activated protein kinase 1; TERT: telomerase reverse transcriptase; NRAS: neuroblastoma RAS viral oncogene homolog; SMARCA4: SWI/SNF related, matrix associated, actin dependent regulator of chromatin, subfamily a, member 4; DICER1: dicer 1, ribonuclease III; NF1: neurofibromin 1; MODY1: Maturity-Onset Diabetes of the Young type 1; RB1: Retinoblastoma 1; AKT1: v-Akt Murine Thymoma Viral Oncogene Homolog 1; NTRAK2: Neurotrophic Receptor Tyrosine Kinase 2; RET: Rearranged during Transfection; IQR: interquartile range; ECOG PS: eastern cooperative oncology group performance status; BM: brain metastasis; NA: not available; PD: progressive disease; SD: stable disease; PR: partial response; CR: complete response; ICI: immune checkpoint inhibitor; PD-1: programmed death-1.

* = Note that some patients had multiple mutations

| Variables | Primary Cohort (N = 152) | |
|---|---|---|
| | *p*-value | HR (95% CI) |
| **Sex (male versus female)** | 0.486 | 0.882 (0.621–1.254) |
| **Smoking status** | | |
| Non-smoker | | 1 |
| Former Smoker | 0.273 | 1.219 (0.865–1.737) |
| Current smoker | 0.774 | 0.948 (0.658–1.365) |
| **Histology (squamous versus non-squamous)** | **<0.001 ***** | 3.170 (1.722–5.837) |
| **Stage at lung cancer diagnosis** | | |
| 1 | | 1 |
| 2 | 0.517 | 0.718 (0.263–1.957) |
| 3 | 0.351 | 1.315 (0.740–2.340) |
| 4 | 0.733 | 1.087 (0.673–1.756) |
| **Location of metastasis at lung cancer diagnosis** | | |
| No metastasis | | 1 |
| Thoracic | **0.007 **** | 1.654 (1.150–2.379) |
| Lymph node | 0.384 | 1.216 (0.783–1.887) |
| Liver | 0.141 | 1.404 (0.894–2.207) |
| Bone | 0.146 | 1.338 (0.904–1.979) |
| **PD-L1 expression** | | |
| Low (<1%) | | 1 |
| Intermediate (1–49%) | 0.743 | 0.938 (0.640–1.375) |
| High (≥50%) | 0.488 | 1.138 (0.790–1.639) |
| Not tested | 0.475 | 1.212 (0.716–2.053) |
| **Mutation status (non-actionable versus actionable)** | **0.034 *** | 0.557 (0.324–0.958) |
| **Primary BM (no versus yes)** | 0.154 | 0.772 (0.541–1.102) |
| **Extracranial metastasis at BM diagnosis (absent versus present)** | 0.090 | 1.758 (0.916–3.374) |
| **Age at BM diagnosis (≥65 versus <65)** | **0.003 **** | 1.767 (1.221–2.555) |
| **ECOG PS at BM diagnosis (≥3 vs 0–2)** | **0.015 *** | 2.477 (1.195–5.133) |
| **Number of BM (>3 versus 1–3)** | **0.021 *** | 1.516 (1.065–2.158) |
| **Largest BM ≥3 cm (no versus yes)** | 0.419 | 1.210 (0.762–1.923) |
| **Clinical benefit to previous line of therapy for BM** | | |

| | | |
|---|---|---|
| No previous treatment | | 1 |
| No clinical benefit | 0.106 | 0.700 (0.455–1.079) |
| Clinical benefit | **0.028 *** | 0.207 (0.051–0.843) |
| **Age at ICI Initiation (≥65 versus <65)** | **0.008 **** | 1.649 (1.140–2.384) |

**Suppl. Table 2: Univariate Cox Proportional Hazards Regression Analysis of Factors Affecting Overall Survival**

Abbreviations: HR: hazard ratio; 95% CI: 95% confidence interval; PD-L1: programmed death-ligand 1; BM: brain metastasis; ECOG PS: eastern cooperative oncology group performance status; ICI: immune checkpoint inhibitor.

* = $p < 0.05$

** = $p < 0.01$

*** = $p < 0.001$

| Regularization models | Area under the curve | Brier score |
|---|---|---|
| Lasso regression | 0.869 | 0.1555 |
| Ridge regression | 0.869 | 0.1574 |
| Elastic Net regression | 0.869 | 0.1571 |
| **Performance metrics** | | **Value** |
| C-index | | 0.630 |
| Brier score | | 0.160 |

**Suppl. Table 3: Comparison of Regularization Models and Performance Metrics for the Internal Validation Model**

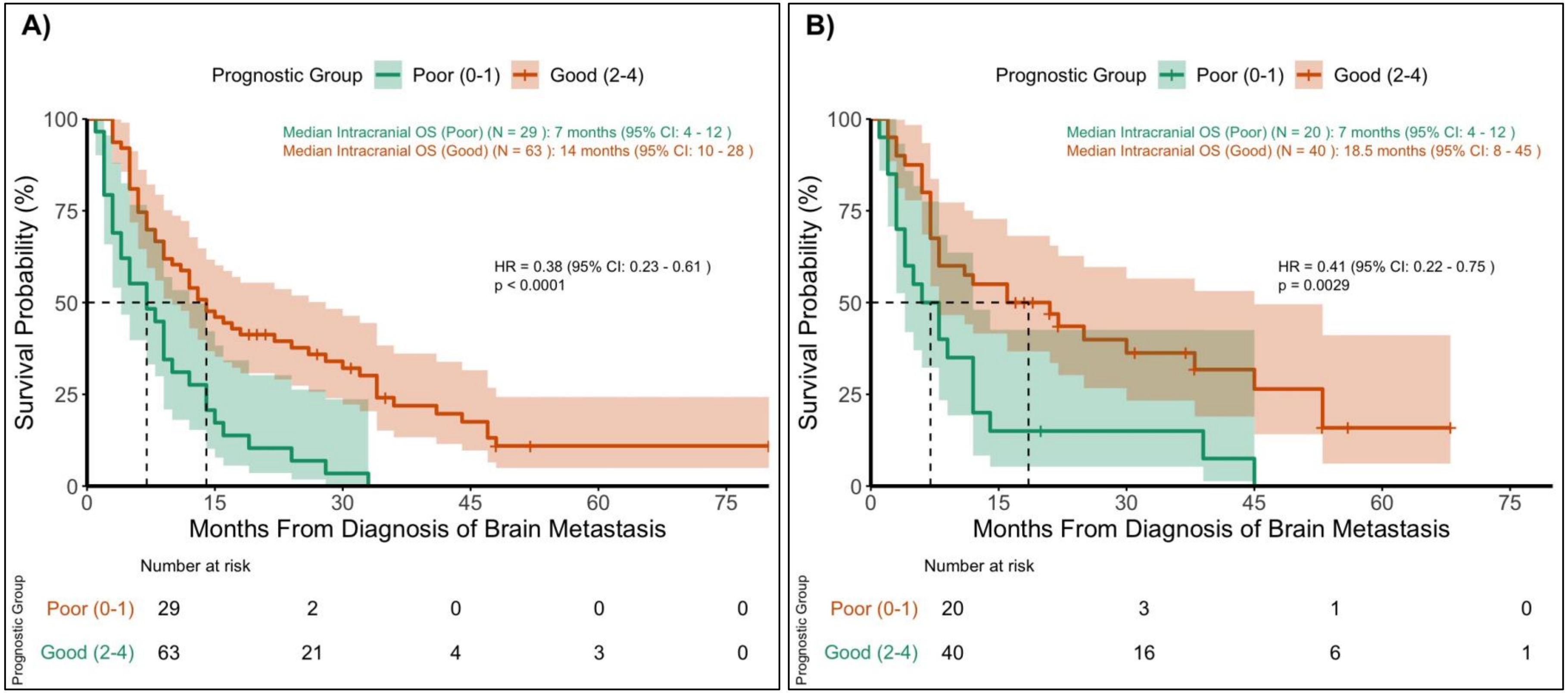


**Suppl. Figure 1: Kaplan-Meier Survival Curves for Prognostic Groups in A) the Training Cohort and B) the Testing Cohort**

Abbreviations: OS: overall survival; 95% CI: 95% confidence interval; HR: hazard ratio.

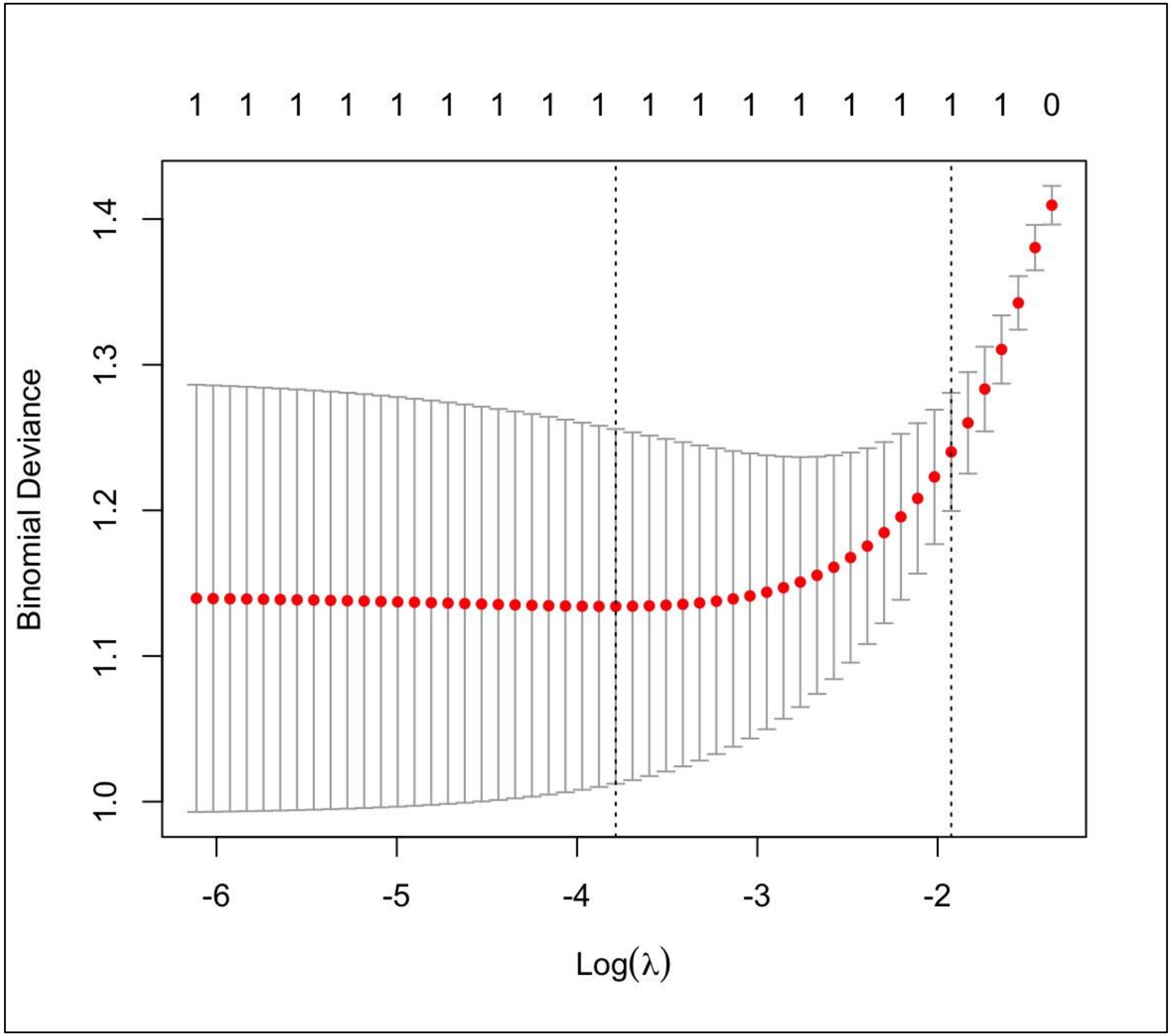


**Suppl. Figure 2: Tuning Parameter (λ) Evaluation Based on Partial Likelihood Deviance with Cross-Validation**
