## Supplementary Data for "The Brain-Lung Immunotherapy Prognostic (BLIP) Score: A Novel Robust Tool for Prognostication in Non-Small Cell Lung Cancer Patients with Brain Metastases"

### Supplementary Data: Methodology

**Data Split**

The dataset was divided into training (60%) and testing (40%) subsets. This approach ensures that the model can be trained on one portion of the data and validated on another, which helps prevent overfitting and provides an unbiased evaluation of the model's performance on unseen data. This data split was used in order to provide a reasonable balance between training and testing data set, considering the limited sample size of the available data with acceptable variance. Kaplan-Meier curves were employed on the training and testing dataset to visualize survival probabilities over time for the different prognostic groups and to determine the feasibility of the data split. The log-rank test was used to compare these survival curves. Additionally, Cox proportional hazards regression was performed to assess the impact of various predictors on intracranial OS and to estimate HRs with 95% CIs.

**Efron-Gong Bootstrap Analysis**

The Efron-Gong Bootstrap method is a sophisticated statistical technique used for the internal validation of predictive models. It addresses the inherent variability in model performance by resampling the data multiple times, thereby providing a robust estimate of the model's accuracy and reliability. This method involves generating multiple bootstrap samples from the original dataset by randomly drawing observations with replacement. Each bootstrap sample is then used to train the predictive model, and the model's performance is evaluated on the out-of-bag (OOB) samples, which are the observations not included in the bootstrap sample for that iteration. The OOB samples serve as a pseudo-test set to assess the model's predictive power. Performance metrics such as accuracy, precision, and recall are calculated for each bootstrap iteration using the OOB samples, resulting in multiple sets of performance metrics. These metrics are then aggregated across all iterations to obtain an overall estimate, typically reported as the mean and standard deviation, summarizing the model's expected performance and its variability. The Efron-Gong Bootstrap method reduces bias by using multiple resampled datasets, provides insights into model stability, and ensures a comprehensive assessment by using OOB samples, making it a preferred choice for a statistically rigorous evaluation of predictive models.

**Penalized Cox Regression and Lasso Regularization Technique**

Penalized Cox regression with Lasso regularization is a robust method for handling high-dimensional survival data, combining the strengths of the Cox proportional hazards model with the variable selection capability of Lasso (Least Absolute Shrinkage and Selection Operator). This approach introduces an $\ell_1$ penalty to the Cox model, which encourages sparsity in the regression coefficients, effectively selecting only the most relevant variables. The objective function minimized in this method is the partial likelihood of the Cox model augmented with an $\ell_1$ penalty, which is expressed as:

$$\mathcal{L}(\beta) = -\sum_{i=1}^{n} \delta_i \left( \beta^T X_i - \log \sum_{j \in \mathcal{R}(t_i)} e^{\beta^T X_j} \right) + \lambda \sum_{j=1}^{p} |\beta_j|$$

where $\delta_i$ is the event indicator, $X_i$ is the covariate vector for the $i$-th individual, ($t_i$) is the risk set at time $t_i$ are the coefficients, $\beta$ are the regression coefficients, and $\lambda$ is the regularization parameter. To determine the optimal value of $\lambda$, we use partial likelihood deviance as the evaluation metric. The deviance is computed as:

$$\text{Deviance}(\lambda) = -2 \log \mathcal{L}(\hat{\beta}(\lambda))$$

where $\hat{\beta}(\lambda)$ are the coefficients estimated for a given $\lambda$. This metric provides a measure of how well the model fits the data. A series of $\lambda$ values are tested to construct a regularization path, which helps in identifying the optimal regularization strength that minimizes the deviance, thereby balancing model fit and complexity.

For internal validation, we employed k-fold cross-validation with $k = 10$. The dataset is partitioned into 10 equally sized folds. In each iteration, 9 folds are used for training the model, and the remaining fold is used for validation. This process is repeated 10 times, with each fold serving as the validation set once. The partial likelihood deviance is computed for each fold, and the average deviance across all folds is used to assess the model's performance. This method ensures that every observation is used for both training and validation, providing a comprehensive evaluation of the model's predictive power and generalizability. The $\lambda$ that results in the lowest average deviance is selected as the optimal regularization parameter, ensuring that the model is neither overfitting nor underfitting the data.

In summary, penalized Cox regression with Lasso regularization, coupled with partial likelihood deviance and k-fold cross-validation, offers a rigorous and effective framework for survival analysis. This method not only improves predictive performance by selecting relevant variables but also ensures model validity and reliability through cross-validation, making it particularly suitable for high-dimensional datasets.

**Performance Metrics**

To assess the performance of our predictive model, we employed two widely recognized metrics: the concordance index (c-index) and the Brier score. These metrics provide complementary insights into the model's discrimination ability and overall prediction accuracy, respectively.

The c-index, also known as the concordance statistic, measures the model's ability to correctly rank the survival times of individuals. It is particularly useful for evaluating models in survival analysis, where the objective is to predict the order of events rather than their exact times. The c-index ranges from 0.5 to 1.0, where 0.5 indicates no better performance than random chance, and 1.0 represents perfect discrimination. A c-index of 1.0 signifies that the model perfectly ranks all pairs of individuals by their predicted risk. In our study, we calculated the c-index by comparing the predicted risk scores against the actual survival times for all possible pairs of subjects. This metric is advantageous because it directly evaluates the model's ability to distinguish between different risk levels in the population.

The Brier score is a measure of the accuracy of probabilistic predictions. Specifically, it calculates the mean squared difference between the predicted probabilities and the actual outcomes, providing a single summary measure of the overall performance. The Brier score ranges from 0 to 1, with lower values indicating better accuracy. A score of 0 indicates perfect prediction, whereas a score of 1 indicates the poorest performance. For our model, we computed the Brier score by comparing the predicted probabilities of survival at specific time points with the observed survival outcomes. The Brier score is particularly useful as it accounts for both the calibration and sharpness of the predicted probabilities, offering a comprehensive evaluation of the model's predictive performance.

By utilizing both the c-index and the Brier score, we obtained a thorough understanding of our model's performance. The c-index provided insights into the model's discrimination power, while the Brier score offered a measure of the overall prediction accuracy. Together, these metrics allowed us to rigorously validate our predictive model, ensuring its reliability and effectiveness in predicting survival outcomes.

**ROC Curve and AUC**

To evaluate the diagnostic performance of our predictive model, we employed Receiver Operating Characteristic (ROC) curve analysis. The ROC curve is a graphical representation that illustrates the diagnostic ability of a binary classifier system as its discrimination threshold is varied. It plots the true positive rate (sensitivity) against the false positive rate (1-specificity) at various threshold settings. The area under the ROC curve (AUC) quantifies the overall ability of the test to discriminate between those individuals with and without the condition. An AUC of 1 indicates perfect discrimination, while an AUC of 0.5 suggests no discriminative power, equivalent to random guessing. For our model, the ROC curve and corresponding AUC were computed to assess the effectiveness of our predictive algorithm in distinguishing between the positive and negative classes

Sensitivity (also known as the true positive rate) measures the proportion of actual positives that are correctly identified by the model, while specificity (the true negative rate) measures the proportion of actual negatives that are correctly identified. These metrics provide a detailed understanding of the model's performance at specific thresholds. High sensitivity ensures that most of the actual positives are correctly detected, which is crucial in contexts where missing a positive case has serious implications. High specificity ensures that most of the actual negatives are correctly identified, reducing the risk of false positives. In our analysis, we calculated sensitivity and specificity at various thresholds to fully understand the trade-offs between these two metrics and to select an optimal threshold that balances them

Youden's J statistic is a summary measure of the ROC curve, used to identify the optimal cut-off point for a diagnostic test. It is defined as:

$$J = \text{Sensitivity} + \text{Specificity} - 1$$

The value of J ranges from 0 to 1, where 1 indicates a perfect test with no false positives or false negatives. By maximizing Youden's J, we can determine the threshold that provides the best balance between sensitivity and specificity. In our study, we calculated Youden's J for different thresholds to identify the point that maximizes the diagnostic accuracy of our model. This statistic is particularly useful because it considers both sensitivity and specificity simultaneously, offering a single value that summarizes the performance of the test.

In summary, the combination of ROC curve analysis, AUC, sensitivity, specificity, and Youden's J statistic provides a comprehensive evaluation of our model's performance. These metrics collectively help in understanding the discriminative power, accuracy, and optimal threshold settings of the predictive model, ensuring its reliability and effectiveness in a clinical setting.

**Decision Curve Analysis**

Decision curve analysis (DCA) is an innovative method employed to evaluate the clinical utility of prognostic scores. It integrates clinical consequences with the statistical performance of the model, providing a comprehensive measure of its practical benefit. The primary advantage of DCA is its ability to weigh the benefits of true positive results against the drawbacks of false positives, considering various threshold probabilities that reflect different clinical scenarios.

In conducting DCA, we first plotted the net benefit for a range of threshold probabilities. The net benefit is defined as the true positive rate minus the false positive rate, adjusted by the ratio of the threshold probability to the probability of a false positive. Mathematically, it can be expressed as:

$$\text{Net Benefit} = \frac{TP}{N} - \frac{FP}{N} \cdot \left( \frac{p_t}{1-p_t} \right)$$

where *TP* represents the number of true positives, *FP* the number of false positives, *N* the total number of patients, and $p_t$ the threshold probability.

To create the decision curves, we calculated the net benefit for each threshold probability and plotted these values. This visual representation allows us to compare the clinical usefulness of different models and strategies, including "treat all" and "treat none" approaches. A model with a higher net benefit across a range of threshold probabilities is considered more clinically valuable.

DCA is particularly beneficial because it does not rely on the prevalence of the outcome, and it accounts for the relative harms of false positives and false negatives in a transparent manner. This makes it an excellent tool for determining whether a predictive model provides a meaningful improvement over standard clinical practice or simpler models. For our study, DCA demonstrated that our predictive model provided superior net benefits compared to existing models across clinically relevant threshold probabilities, underscoring its potential utility in improving patient outcomes.

By employing DCA, we ensured that our model's validation was not only statistically sound but also clinically relevant, supporting its implementation in real-world decision-making processes.
